# Impact of the COVID-19 pandemic on antidiabetic drugs initiation in Sweden, an interrupted time series study

**DOI:** 10.1101/2024.05.09.24307137

**Authors:** Nataliia Khanyk, Fredrik Nyberg, Björn Wettermark, Huiqi Li, Katarina Eeg-Olofsson, Soffia Gudbjornsdottir, Mohammadhossein Hajiebrahimi

## Abstract

**Aim:** To investigate the impact of the COVID-19-pandemic on antidiabetic drug initiation

**Methods:** Using Swedish national health registers, an interrupted time series study for all first-time dispensed antidiabetic drugs was performed. A minimum 14-month washout period was used. We calculated monthly cumulative incidence of antidiabetic drug initiations per 1000 population and estimated trends before and after the onset of the pandemic using segmented regression analysis.

**Results:** Among 167,889 participants between 1Mar2019 and 30Nov2021, male sex (53%), age 18-64 (55.3%), unmarried status (52.5%), upper secondary education level (46.2%), and being employed (53.5%) were the most frequent characteristics. Mar2020, there was an immediate level drop in antidiabetic drug initiation (β_2=_-0.087%, P=0.01); however, the trend exhibited a rapid increase thereafter. Significant immediate level drops were identified for DPP4i (−0.087%, P=0.01), GLP1a (−0.008%, P=0.01) and SGLT2i (−0.033%, P=0.003). Older people (≥65) showed the largest immediate drop for antidiabetic drug initiation (−0.272%, P=0.003). SGLT2i demonstrated an increasing pattern both before and after the pandemic while metformin did reveal an increasing pattern among patients ≥65 years old after the pandemic.

**Conclusion:** The results identified an immediate level drop at the onset of the pandemic for any antidiabetic drug initiation, but the trend increased rapidly thereafter.

## 1. Introduction

The impact of severe acute respiratory syndrome coronavirus 2 (SARS-CoV-2) and the associated infection named Coronavirus Disease 2019 (COVID-19) on patients with type 2 diabetes mellitus (DM2) is an important clinical challenge for both patients [1] and health services. It has been shown that COVID-19 increases the risk of new onset of DM2 [2], and conversely, pre-existing DM2 confers an increased risk of severe COVID-19, increasing the risk of hospitalization and death [3–9].

Association between particular antidiabetic drugs and COVID-19 infection or severity has been considered in some studies. Metformin, glucagon-like peptide-1 analogues (GLP1a), and sodium glucose co-transporter 2 inhibitors (SGLT2i) were found to be associated with reduced risk of COVID-19 mortality [10–12]. Insulin use might increase the risk of mortality in persons with diabetes with COVID-19 [10,13–16]. Several reviews have suggested that the effect of antidiabetic agents in patients with DM2 and COVID-19 requires further exploration, and RCTs are needed [14,17,18].

Regarding the effect of the pandemic on initiation of antidiabetic drugs, there are few studies and most indicate some significant impact on drug use [19–21]. A considerable decrease in the number of weekly insulin prescription fills was seen during, compared to before, the COVID-19 pandemic in the US (January 2019 and October 2020) [21], and lower rate of treatment initiations with antidiabetic drugs during lockdown and an increasing rate in treatment disruptions afterward was reported in France [20]. Furthermore, a temporary but statistically significant immediately decrease in antidiabetic drugs initiation at the beginning of the pandemic, followed by a significant increase in slope, was reported in Canada during the first and second quarters of the pandemic [19].

Given the relative scarcity of investigations on the impact of the COVID-19 pandemic on antidiabetic drug initiation, especially on specific antidiabetic drug classes, we aimed to examine the initiation of antidiabetic drugs in Sweden before and during the COVID-19 pandemic.

## 2. Materials and Methods

An interrupted time series (ITS) study was performed to evaluate changes in the incidence of new antidiabetic drug use in Sweden between 1 Jan 2018 and 30 November 2021. March 2020 was selected as the interruption point to compare trend changes before and following the onset of the pandemic. Using Anatomical Therapeutic Chemical (ATC) codes, the first prescription of an antidiabetic drug was studied, and a minimum prior washout period without antidiabetic use was applied, see 3.1. Five different classes of antidiabetic drugs were included in the analysis: insulins and analogues (ATC code A10A), biguanide derivatives (A10BA), glucagon-like peptide-1 analogue (GLP-1a) (A10BJ), sodium-glucose co-transporter 2 inhibitors (SGLT-2i) (A10BK), and dipeptidyl peptidase 4 inhibitors (DPP4i) (A10BH) (Supplementary Table 1). Antidiabetic drug initiators who died or emigrated during each time period were included in that period but excluded from the next period, so that people who were alive and lived in Sweden at the beginning of each month were the denominator population for that month.

This study is nested in and used data from the Swedish Covid-19 Investigation for Future Insights – a Population Epidemiology Approach using Register Linkage (SCIFI-PEARL) project database [22], which now includes the entire population in Sweden linked with rich health-related and socioeconomic data, including all positive PCR tests for COVID-19. Linkage was performed using the Swedish personal identification number [23], and data are available in pseudonymized form for the SCIFI-PEARL project. Data was collected at the event time by the registers and SCIFI-PEARL received this data every 3 months. Data from the following registers were used for this study:

- The National database of notifiable diseases (SmiNet) [24] – collects data on SARS-Cov-2 test positive results since the beginning of the pandemic.
- National Prescribed Drug Register [25] – collects data for all dispensed prescription drugs in Sweden. Data are available in SCIFI-PEARL from 1 Jan 2018.
- National Patient Register [26] – collects data on inpatient and specialist outpatient care. Data are available in SCIFI-PEARL from 1 Jan 2015.
- LISA: Longitudinal Integrated database for Health Insurance and Labour Market Studies at Statistics Sweden – collects data on sociodemographic factors for all persons registered in Sweden [27].
- Total population register [28] – collects data on migration and population births and deaths.
- National Cause of Death Register [29] – collects data on all deaths including date and cause of death.

The patients dispensed antidiabetic drugs were characterized by sex (male, female), age group (<18, 18-64, and ≥65 years old), marital status (married, unmarried), educational level (up to end of lower secondary school (≤9 years), up to end of secondary school (≤12 years), tertiary school and academic (>12 years)), country of birth (Sweden, Nordics excluding Sweden, EU28 except the Nordics, Out of EU28), and employment status (unemployed including retired, employed). Furthermore, the number of different drugs dispensed other than antidiabetic drugs (0, 1, 2, 3-4, 5-9, 10-19, ≥20) before the start of the study period (between 1 Jan 2018, and 28 Feb 2019), as well as the number of comorbidities (1, 2, 3,4, ≥5) between 1 Jan 2015 and 28 Feb 2019 were calculated for all antidiabetic drug initiators in the study. We also identified the ten most frequent other non-antidiabetic drugs dispensed and ten most frequent comorbidities before the pandemic.

### 2.1 Statistical methods

Descriptive statistics were used to present frequency and percentage of characteristics of participants of the study. Monthly cumulative incidence rates were calculated for all new antidiabetic drug use in those that were not taking any antidiabetic drugs during the washout period. The washout period was defined as a fixed period between 1 Jan 2018 and 28 Feb 2019. Since the first prescription date for any individual has been used as their index date, the washout period will be longer for later months in the study. Cumulative incidence rates of new initiation were also calculated in the same way for new users of each separate class of antidiabetics when they were naïve for that particular antidiabetic class. Based on this definition, the study population differs for different antidiabetic drug classes, and we observe different characteristics of antidiabetic users in the study for the particular drug classes (Supplementary Table 2).

A single interrupted time series (SITSA), using a segmented regression model, estimated monthly cumulative incidence of antidiabetic drugs initiations per 1000 population [30]. Trends of antidiabetic drug initiation before and after onset of the pandemic and immediate or late trend changes were estimated. Twelve time points before the pandemic and 20 time points after the onset of the pandemic were considered in the study. The single interrupted time series analysis using segmented regression model and their contexts have been described in detail elsewhere [30]. In brief, the equation of the model was specified as follows: y= α + β_1_T + β_2_X+ β_3_XT+ε where y = outcome variable, α = intercept, β = coefficients (β_1_= pre-intervention trend, β_2_=level change following the intervention, β_3_= post-intervention trend, and thus β_1_+ β_3_=post intervention slope), T= time, X = study phase, XT= time after the interruption, and ε = error or residual.

All analyses were performed using SAS 9.4. The research has ethics approval from the Swedish Ethical Review Authority (2020-01800 with subsequent amendments).

## 3. Results

The study included 167,889 individuals who initiated certain antidiabetic drugs between 1 March 2019 and 30 November 2021. Table 1 demonstrates the baseline characteristics of the participants. The median age was 62 years (IQR: 49-72). The majority of antidiabetic drug new users were born in Sweden (71.1%) while more than one fifth (21.3%) were born in countries outside the EU28. Male sex (53.0%), age 18-64 (55.3%), unmarried status (52.5%), upper secondary education level (46.2%), and being employed (53.5%), were the most frequent characteristics among the new antidiabetic drug users.

**Table 1.** Characteristics of patients with no history of antidiabetic drug use since 1 Jan 2018, at their first dispensing of any antidiabetic drug in Sweden from 1 March 2019 to 30 Nov 2021.

| **<u>Variables</u>** | **<u>Numbers</u>** | **<u>Percent</u>** | **<u>P-value</u>** |
| --- | --- | --- | --- |
| **<u>Total</u>** | 167,889 |  |  |
| **<u>Sex</u>** |  |  |  |
| Male | 89,020 | 53.0 | <0.0001 |
| Female | 78,869 | 47.0 |  |
| **<u>Year at diagnosis</u>** |  |  |  |
| 1 March 2019-28 Feb 2020 | 52,510 | 31.3 | <0.0001 |
| 1 March 2020-28 Feb 2021 | 54,777 | 32.6 |  |
| 1 March 2021-30 Nov 2020 | 60,602 | 36.1 |  |
| **<u>Age at January 2020</u>** |  |  |  |
| <18 | 2,466 | 1.5 | <0.0001 |
| 18-64 | 92,781 | 55.3 |  |
| ≥65 | 72,642 | 43.3 |  |
| Median (IQR) | 62 (49-72) |  |  |
| Range | 6-103 |  |  |
| **<u>Marital Status at January 2020</u>** |  |  |  |
| Married | 79,471 | 47.5 | <0.0001 |
| Unmarried | 87,796 | 52.5 |  |
| Missing | 622 |  |  |
| **<u>Educational level at January 2020</u>** |  |  |  |
| Up to end of secondary school (9 years) | 42,505 | 26.6 | <0.0001 |
| Up to end of upper secondary school (12 years) | 73,823 | 46.2 |  |
| Tertiary school and Academic (More than 12 years) | 43,632 | 27.3 |  |
| Missing | 7,929 |  |  |
| **<u>Occupation status at January 2020</u>** |  |  | <0.0001 |
| Employed | 87,932 | 53.5 |  |
| Unemployed/Retired | 76,440 | 46.5 |  |
| Missing | 3,517 |  |  |
| **<u>Country of birth</u>** |  |  |  |
| Sweden | 119,335 | 71.1 | <0.0001 |
| Nordics excluding Sweden | 6,192 | 3.7 |  |
| Eu28 except the Nordics | 6,517 | 3.9 |  |
| Out of Eu28 | 35,824 | 21.3 |  |
| Missing | 21 |  |  |

All studied types of initiated antidiabetic drugs are shown in Table 2. Overall, 91.4% of users had only one antidiabetic drug prescribed at their first visit with a new antidiabetic initiation. Metformin was the most commonly newly prescribed antidiabetic drug, found in three quarters of all study participants (75.5%) followed by insulins (10.7%), GLP-1a (5.0%), SGLT2i (4.9%), and DPP4i (2.8%). The characteristics of the study population for new initiators of each specific antidiabetic drug class are presented in Supplementary Table 2. DPP4 inhibitors were most often dispensed to new users aged over 65 (75.5%) and unemployed or retired patients (71.5%). SGLT2 inhibitors were more frequent among males (69.0%) and also among those above 65 years of age (58.0%). GLP-1a, in contrast, were most frequently initiated by patients in the 18-64 age group (84.1%) and by females (66.4%).

**Table 2:**
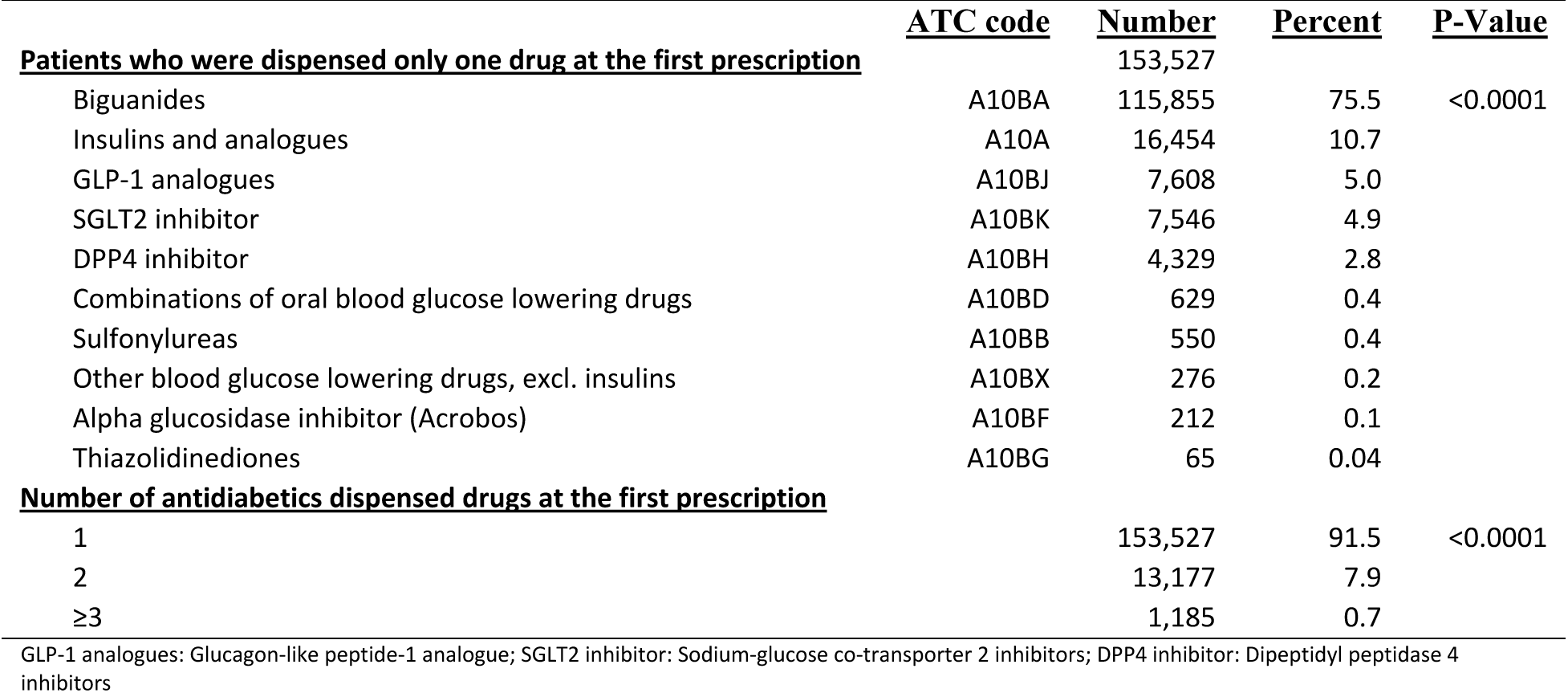
Type of antidiabetic drugs dispensed to patients with no prior antidiabetic drug history since 1 Jan 2018, at their first dispensing of any antidiabetic drug in Sweden from 1 March 2019 to 30 Nov 2021.

The number of different non-antidiabetic drug substances prescribed and number of comorbidities in the study population registered in specialist out- and inpatient care during the years before the study period are shown in Table 3. Among all antidiabetic drug users, 32.9% were dispensed 5-9 non-antidiabetics drugs and 19.0% had 3-4 drugs in the 2-year period before the study period, together more than half of all study participants. Regarding comorbidities in the 4 years and 2 months preceding the study period, more than half of participants had more than 3 comorbidity diagnoses recorded in hospital out- or inpatient care.

**Table 3:**
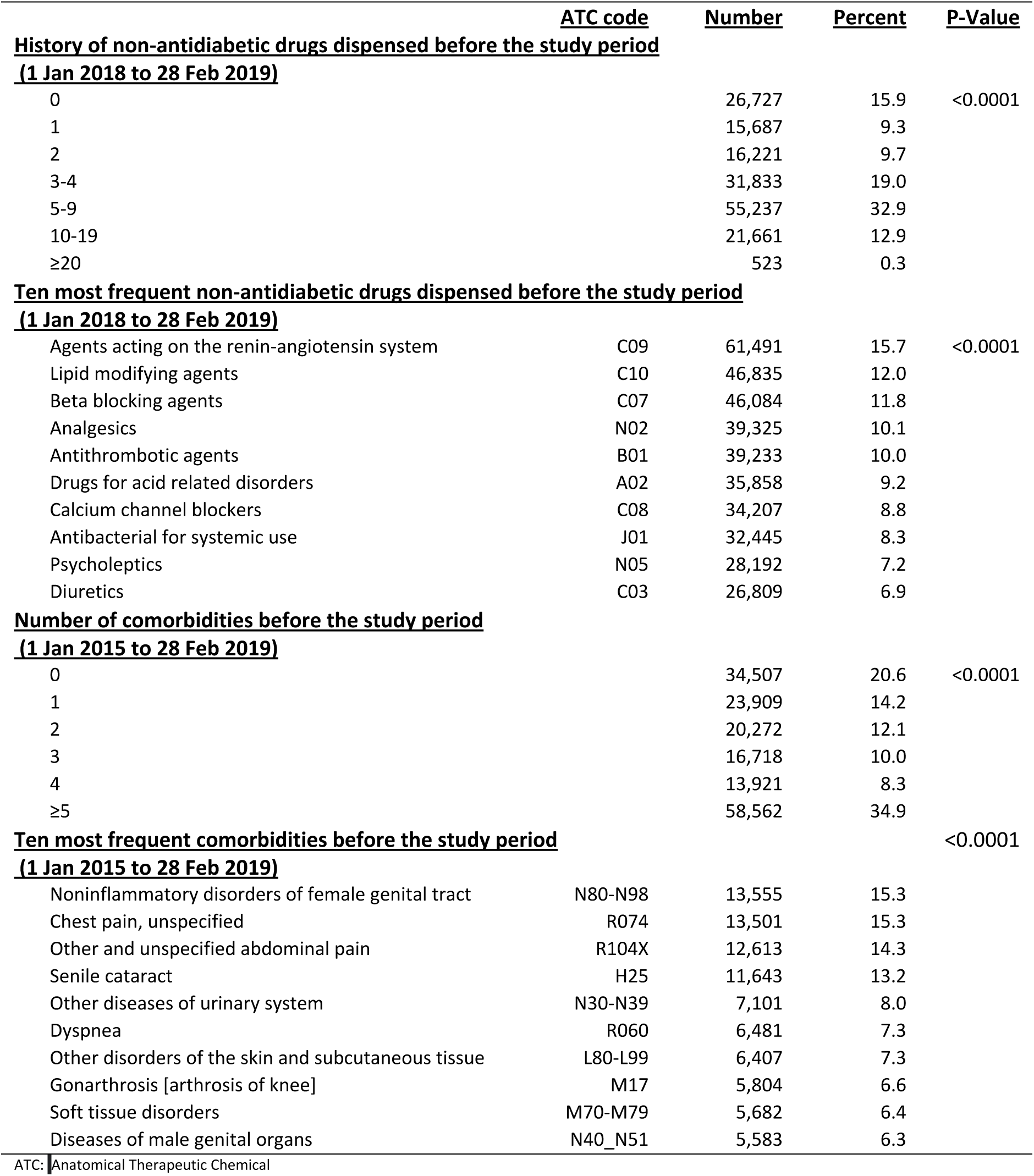
Distribution of non-antidiabetic drug dispensing and comorbidities other than diabetes prior to the start of the study period (March 2019), among new users of antidiabetic drugs in Sweden 1 March 2019 to 30 Nov 2021.

The results of the ITS analysis estimated an immediate significant level change in antidiabetic drug initiation following the onset of the pandemic, with the rate (ß2) dropping by −0.087% between the pre-and post-pandemic periods for all antidiabetic drugs. A significant level change was identified for the specific drug classes of DPP4 inhibitors (ß2 = −0.087%, P=0.010) (Figure 1A and Supplementary Table 3a), GLP1 analogues (ß2 = −0.008%, P=0.014) (Figure 1c and Supplementary Table 3c) and SGLT2 inhibitor (ß2 = −0.033%, P=0.003) (Figures 1C and Supplementary Table 3c). We did not find a significant immediate change at the start of the pandemic when we considered metformin initiation (ß2 =-0.036, P=0.224) or insulin treatment regardless of its indication (ß2 =0.002, P=0.609). New antidiabetic drug users over 65 years old showed a stronger immediate level change, with the rate decreasing by −0.272% (P=0.003) for all antidiabetic drugs (Figure 1A and Supplementary Table 3a) and −0.122% for SGLT2 inhibitors (P=0.006) at the beginning of the pandemic (Figure 1C and Supplementary Table 3c).

**Figure 1A:**
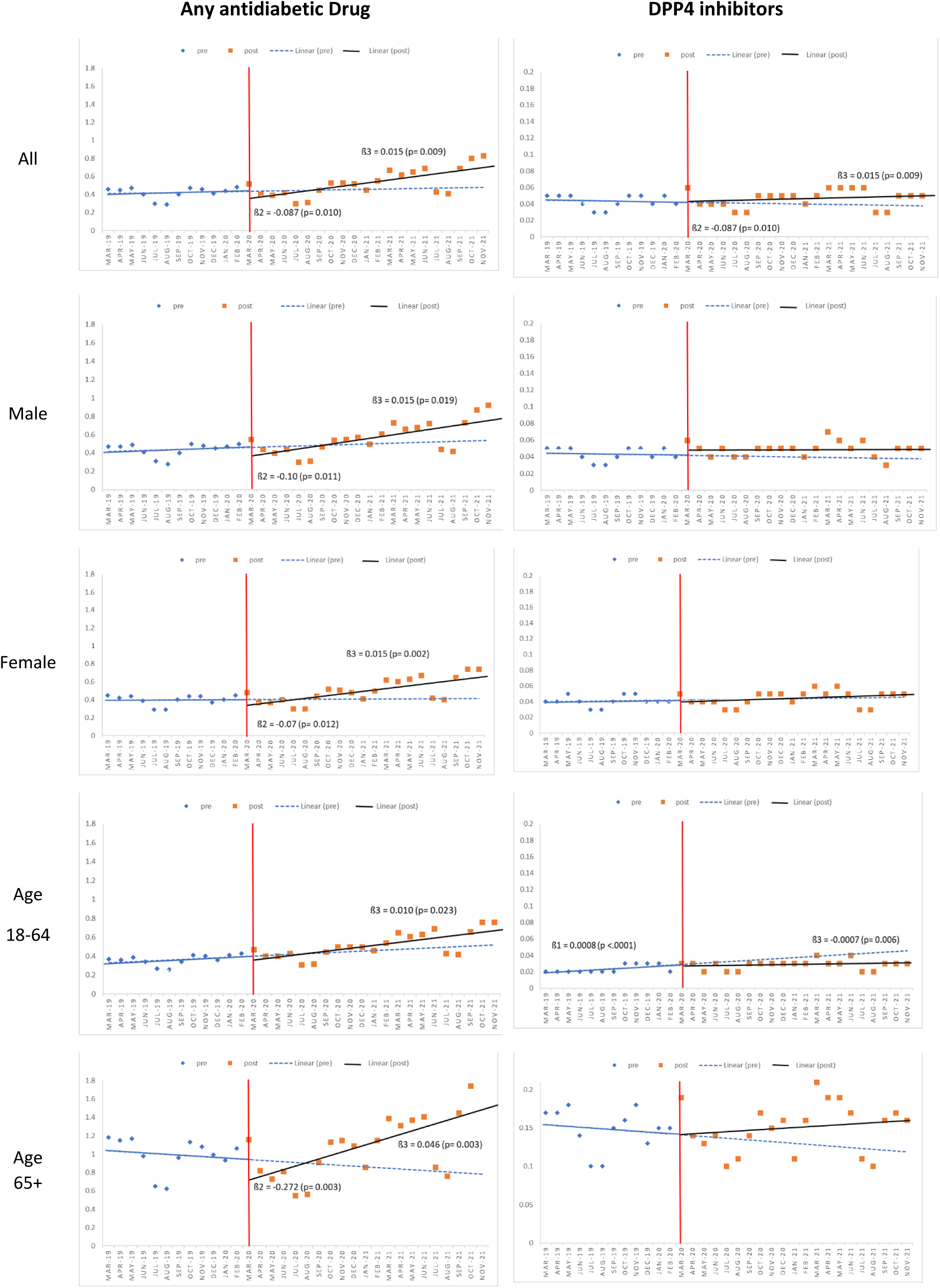
Linear segmented regression analysis for cumulative incidence rate of initiating any antidiabetic drug (left), and DPP4 inhibitors (right) before and after the pandemic in Sweden. (Only statistically significant results shown); DPP4 inhibitor: Dipeptidyl peptidase 4 inhibitors

Regarding the long-term trend of antidiabetic drug dispensing, initiation of all antidiabetic drugs (Figure 1A and Supplementary Tables 3a), SGLT2 inhibitors and GLP1 inhibitors showed similar pattern after the immediate change compared with the trend before the pandemic (Figure 1C and Supplementary Tables 3c). However, for metformin (among females and patients aged ≥65 years) (Figure 1B and Supplementary Table 3b) and for insulin (among all users, females and patients aged ≥65 years) (Figure 1B and Supplementary Table 3b), the initiation pattern was changed from a decreasing trend before the pandemic to an increasing pattern after the pandemic. Moreover, the initiation pattern of DPP4 inhibitors also changed from decreasing before the pandemic to increasing after the pandemic among patients at the age 18-64.

**Fig. 1B:**
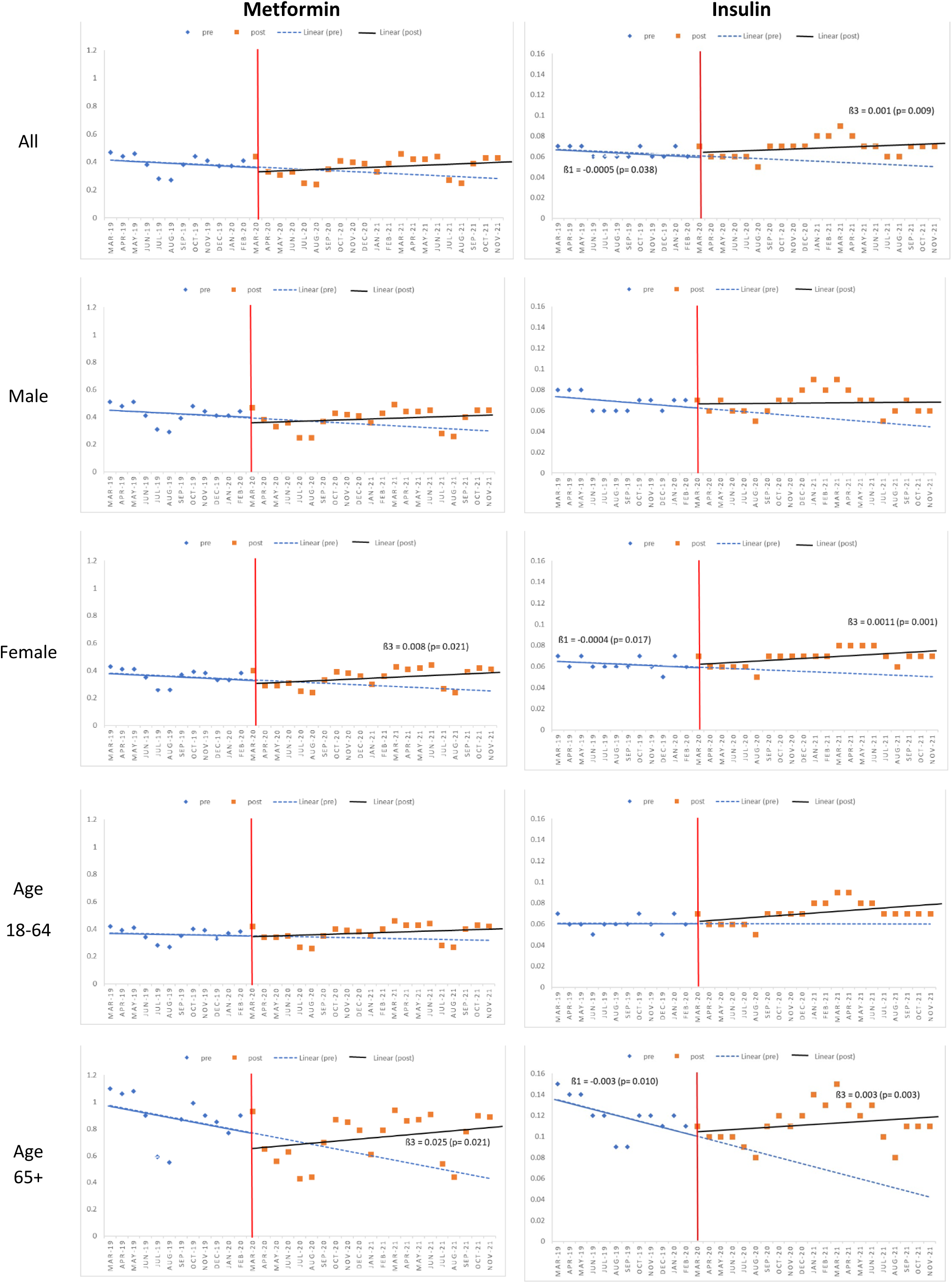
Linear segmented regression analysis for cumulative incidence rate of initiating metformin (left) and insulin (right) before and after the pandemic in Sweden, March 2019 to Nov 2021. (Only statistically significant results shown)

**Fig. 1C:**
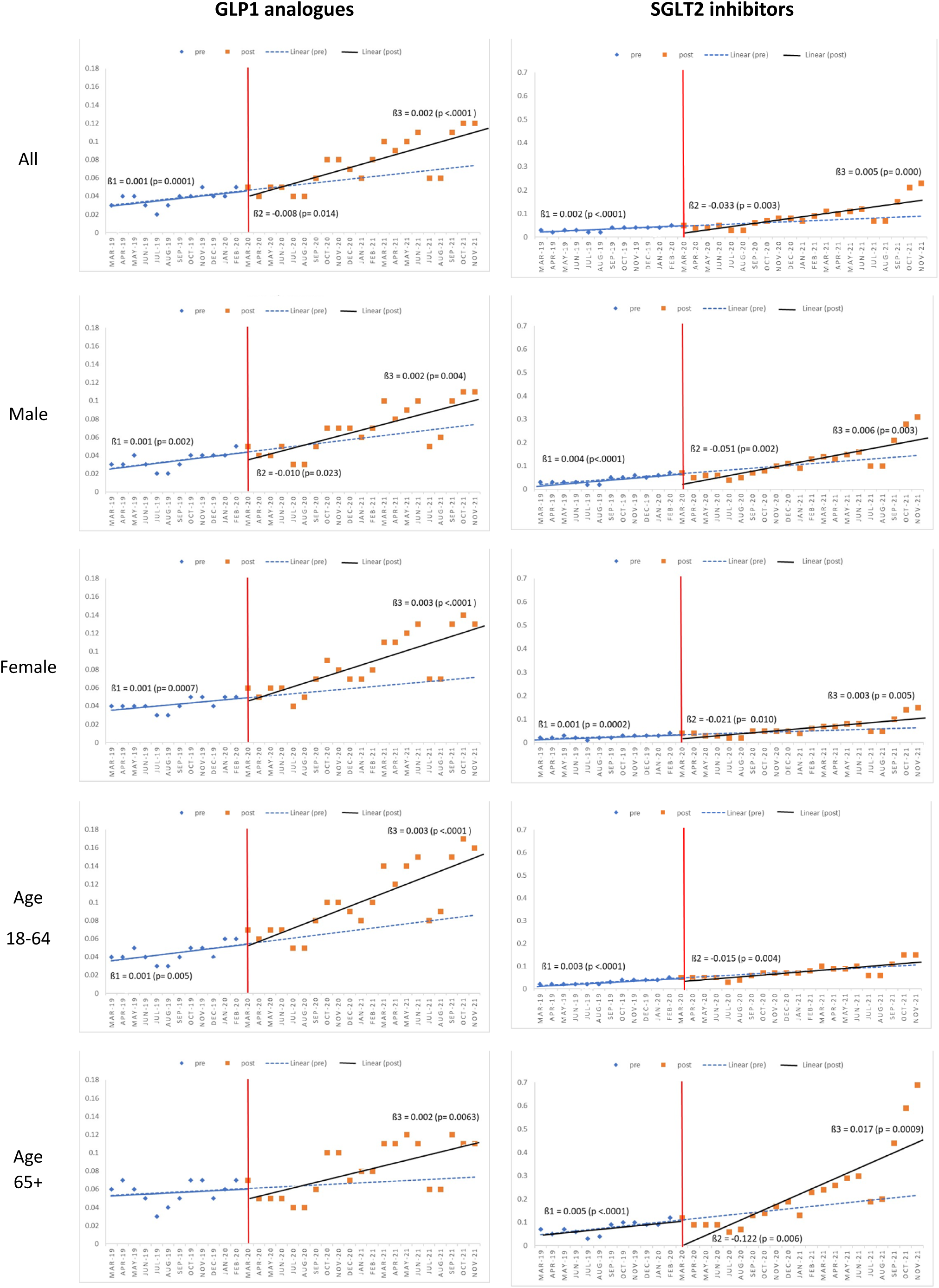
Linear segmented regression analysis for cumulative incidence rate of initiating GLP1 analogues (left) and SGLT2 inhibitors (right) before and after the pandemic in Sweden, March 2019 to Nov 2021. (Only statistically significant results shown); DPP4 inhibitor: Dipeptidyl peptidase 4 inhibitors

## 4. Discussion

In this ITS study, an immediate level drop in incidence of all antidiabetic drugs was observed at initiation of the pandemic. The level drop was notable for both sexes and stronger among patients aged ≥65 years. The most substantial immediate level drop occurred within the class of SGLT2 inhibitors. The level drop in SGLT2 inhibitors use was observed across both sexes and all age subgroups, but with the most prominent effect in the older population.

Considering all antidiabetic drugs, the results revealed a substantial level drop in drug initiation at the onset of the pandemic, although not statistically significant for people aged 18-64 years. This observation may suggest appropriate adherence of Swedes to the recommendations aimed to minimizing interactions during the pandemic, despite the relatively unique approach of Sweden in tackling the pandemic. Sweden did not implement a lockdown, obligatory facemasks outside healthcare, or quarantines for infected households or geographical regions, and did not close kindergartens and schools for children up to 16 during the COVID-19 pandemic. Instead, Sweden adopted various measures with a focus on recommendations such as enhanced hygiene practices, general distance-working for employed, physical distancing in public spaces and banned visits to nursing facilities to avoid unnecessary healthcare visits and close contacts particularly for people aged 70 and above [31,32]. The results of this study might reflect this strategy in the country.

Furthermore, it was observed that the drop in the drug initiation for all antidiabetic groups at the beginning of the pandemic onset was more prominent among elderly individuals. This likely mirrors the higher vulnerability of this age group to COVID-19, where age stands out possibly as the most important factor influencing the severity of COVID-19, as was identified early on [33,34]. Moreover, the persons with diabetes were considered a risk group and advised to avoid contact with others. Therefore, patients aged over 65 may have been reluctant to locations with the potential for larger population, including healthcare and pharmacy locations, and may prefer to consult with healthcare professionals exclusively in urgent situations.

The findings also demonstrated specifically an immediate level drop in the incidence of new SGLT2 inhibitors drug dispensing at the onset of the pandemic. However, the utilization of SGLT2 inhibitors increased substantially before and during the pandemic. While the initial change in level at the onset of pandemic could be attributed to the Swedish pandemic strategy, the rise in utilization is likely associated to the new indication of the SGLT2 inhibitors for reducing cardiovascular risk in patients with diabetes [11,35,36]. The sharp increase in the trend of SGLT2 inhibitors among people aged 65 and older likely reflects the influence of these new indications [37].

The results indicated no substantial changes for metformin, despite an increasing pattern of initiation during the pandemic, particularly among individuals aged over 65. However, there was a notable increasing pattern in GLP1 dispensing, although not statistically significant, observed for both sexes and age subgroups. Similar to the SGLT2 inhibitors group, the increasing pattern seen for GLP1 analogues might be attributed to the new indication of the drug for weight reduction [38,39] and the reduction of cardiovascular risk among patients with DM2 [11,36].

Our study has several strengths including its comprehensive coverage of the Swedish population and its ability to integrate various data sources from different health registries using unique personal ID numbers. We, in fact, were thus able to include all dispensed antidiabetic drugs in Sweden in the study. Our study had some limitations as well. The Swedish Prescribed Drug Register does not include the indication of the prescribed drugs. Therefore, it was not possible to determine the main indication for prescription of a particular drug, if there were several indications for that particular drug. Additionally, for the sub-analyses of specific antidiabetic drugs in this study, the term “new users” in the context of separate antidiabetic classes refers to individuals who were new users within their respective class but not necessarily new users of any antidiabetic medication as a whole. We also used a fixed 14-month pre-study washout period, which gives a longer washout period for later participants in the study. The 14-month was the period between 2018.01.01 (the first available drug data date in PDR) and 1 mar 2020 (the pandemic month), however, the later participants had a longer washout period. Although participants did not have the same washout period in the study, it helped us to avoid having a participants two times in the study if she/he stopped using antidiabetic for 14 months and started it again. Finally, recorded diagnoses and comorbidities were captured from the National Patent Register, which does not include primary care.

In conclusion, an immediate drop changes occurred at the onset of the pandemic, followed by a rapid return to comparable or in some instances, increased level a few months later, with the elderly population exhibiting the most pronounce changes. This pattern could be interpreted as an outcome of the pandemic-related restrictions and recommendations introduced initially in Sweden, leading to a transient reduction in visits to healthcare institution as well as pharmacies.

## Funding

This work was supported by the SCIFI-PEARL project which has basic funding based on grants from the Swedish state under the agreement between the Swedish government and the county councils, the ALF-agreement (Avtal om Läkarutbildning och Forskning/Medical Training and Research Agreement) grants ALFGBG-938453, ALFGBG-971130, ALFGBG-978954 and previously from a joint grant from Forte (Swedish Research Council for Health, Working Life and Welfare) and FORMAS (Forskningsrådet för miljö, areella näringar och samhällsbyggande), grant 2020-02828., as well as a grant from the Swedish Heart-Lung Foundation (20210581). This work also was supported by the Nordic COHERENCE Project, Project No. 105670 funded by NordForsk under the Nordic Council of Ministers.

## Authors’ relationships and activities

We declare no competing interests.

## Data Availability

The data underlying this article cannot be shared publicly due to their containing sensitive information that could compromise the privacy of research participants. Analysis data will be shared upon reasonable request to the authors. Access to similar data requires permission. Apart from ethical approval from the Swedish Ethical Review Authority, researchers will also need approval from each register holder.

## Author contributions

MH, and BW wrote the protocol and designed the study. FN and BW were responsible for ethics and obtaining data. FN and HL were responsible for data linkage and compilation and basic data quality control and data management. MH has prepared the first draft of the results. MH conducted the analysis-specific data management, and statistical analyses. NK and MH prepared the first draft of the manuscript. All authors (including KEO, SG) have critically participated in reviewing and interpreting of the results and revising the manuscript. All authors have approved the final manuscript.

## Declaration of Competing Interest

KEO has received fees for lecturing and/or honoraria for consulting from Sanofi, Novo Nordisk, Eli Lilly and Abbot Diabetes Care. FN has some AstraZeneca shares. The remaining authors have no known competing financial interests or personal relationships that could have appeared to influence the work reported in this paper.

## Acknowledgments

The authors acknowledge the initial work of former master student in pharmacy Nicole Kastrati, conducted at Uppsala University (supervised by MH). The authors also acknowledge to the Nordic COHERENCE Project for supporting our study and the Wenner-Gren foundation (GFU2022-0008) for providing financial support to NK.

## Supplementary materials

**Supplementary Table 1:** ATC codes for antidiabetic drugs included in the study.

| **<u>Groups</u>** | **<u>ATC codes</u>** |
| --- | --- |
| **Insulin and analogues** | **A10A** |
| **Blood glucose-lowering agents excluding Insulin** | **A10B** |
| **Biguanide derivatives** | **A10BA** |
| Metformin | A10BA02 |
| **Sulfonylurea compounds** | **A10BB** |
| Glibenclamide | A10BB01 |
| **Peroral diabetes drugs, combinations** | **A10BD** |
| **Glycosidase inhibitors** | **A10BF** |
| Acarbose | A10BF01 |
| **Thiazolidinediones** | **A10BG** |
| **Dipeptidyl peptidase 4 inhibitors (DPP-4i)** | **A10BH** |
| **Glucagon-like peptide-1 analogue (GLP-1a)** | **A10BJ** |
| **Sodium-glucose co-transporter 2 inhibitors (SGLT2i)** | **A10BK** |
| **Other blood glucose lowering agents excluding Insulin** | **A10BX** |

**Supplementary Table 2.** Characteristics of the new users of antidiabetic drugs by type of drug, in Sweden March 2019 to the end of November 2021.

| <u>Variables</u> | <u>Insulin</u> |  | <u>Metformin</u> |  | <u>DPP4i</u> |  | <u>GLP1a</u> |  | <u>SGLT2i</u> |  |
| --- | --- | --- | --- | --- | --- | --- | --- | --- | --- | --- |
|  | <u>Number</u> | <u>Percent</u> | <u>Numbers</u> | <u>Percent</u> | <u>Numbers</u> | <u>Percent</u> | <u>Numbers</u> | <u>Percent</u> | <u>Numbers</u> | <u>Percent</u> |
| **<u>Total</u>** | 26,530 |  | 112,158 |  | 5,650 |  | 9,161 |  | 10,234 |  |
| **<u>Sex</u>** |  |  |  |  |  |  |  |  |  |  |
| Male | 13,887 | 52.34 | 59,606 | 53.14 | 2,850 | 50.44 | 3,090 | 33.73 | 7,059 | 68.98 |
| Female | 12,643 | 47.66 | 52,552 | 46.86 | 2,800 | 49.56 | 6,071 | 66.27 | 3,175 | 31.02 |
| **<u>Age at the first dispensing date</u>** |  |  |  |  |  |  |  |  |  |  |
| <18 | 1,739 | 6.55 | 668 | 0.60 | 3 | 0.05 | 62 | 0.68 | 9 | 0.09 |
| 18-64 | 13,198 | 49.75 | 64,043 | 57.10 | 1,379 | 24.41 | 7,709 | 84.15 | 4,286 | 41.88 |
| ≥65 | 11,593 | 43.70 | 47,447 | 42.30 | 4,268 | 75.54 | 1,390 | 15.17 | 5,939 | 58.03 |
| **<u>Marital Status at the pandemic</u>** |  |  |  |  |  |  |  |  |  |  |
| Married | 10,331 | 39.20 | 55,199 | 49.36 | 2,421 | 43.06 | 4,271 | 46.70 | 5,162 | 50.61 |
| Unmarried | 16,026 | 60.80 | 56,632 | 50.64 | 3,202 | 56.94 | 4,874 | 53.30 | 5,037 | 49.39 |
| Missing | 173 |  | 327 |  | 27 |  | 16 |  | 35 |  |
| **<u>Educational level at the pandemic</u>** |  |  |  |  |  |  |  |  |  |  |
| Secondary school (9 years) | 7,206 | 30.31 | 27,956 | 25.87 | 2,055 | 37.64 | 1,500 | 16.74 | 2,694 | 27.06 |
| Upper secondary school (12 years) | 10,554 | 44.39 | 50,522 | 46.76 | 2,289 | 41.93 | 4,096 | 45.70 | 4,671 | 46.92 |
| Academic education (>12 years) | 6,018 | 25.31 | 29,574 | 27.37 | 1,115 | 20.42 | 3,367 | 37.57 | 2,591 | 26.02 |
| Missing | 2,752 |  | 4,106 |  | 191 |  | 198 |  | 278 |  |
| **<u>Occupation status at the pandemic</u>** |  |  |  |  |  |  |  |  |  |  |
| Employed | 11,376 | 45.98 | 61,306 | 55.32 | 1,592 | 28.50 | 6,835 | 75.15 | 4,796 | 47.35 |
| Unemployed/Retired | 13,367 | 54.02 | 49,514 | 44.68 | 3,993 | 71.50 | 2,260 | 24.85 | 5,333 | 52.65 |
| Missing | 1,787 |  | 1,338 |  | 65 |  | 66 |  | 105 |  |
| **<u>Country of birth</u>** |  |  |  |  |  |  |  |  |  |  |
| Sweden | 19,600 | 73.90 | 78,215 | 69.74 | 4,575 | 80.99 | 6,239 | 68.13 | 8,200 | 80.13 |
| Nordics excluding Sweden | 984 | 3.71 | 4,064 | 3.62 | 249 | 4.41 | 282 | 3.08 | 464 | 4.53 |
| Eu28 except the Nordics | 966 | 3.64 | 4,328 | 3.86 | 199 | 3.52 | 459 | 5.01 | 367 | 3.59 |
| Out of Europe | 4,974 | 18.75 | 25,541 | 22.77 | 626 | 11.08 | 2,178 | 23.78 | 8,200 | 80.13 |
| Missing | 6 |  | 10 |  | 1 |  | 3 |  | 1 |  |

GLP-1 analogues: Glucagon-like peptide-1 analogue; SGLT2 inhibitor: Sodium-glucose co-transporter 2 inhibitors; DPP4 inhibitor: Dipeptidyl peptidase 4 inhibitors

**Supplementary Table 3a:** Single ITS analysis (SITSA) for cumulative incidence rate of any antidiabetic drug and DPP4i initiation before and after the pandemic in Sweden.

| Antidiabetic drugs |  |  |  | DPP4i |  |  |
| --- | --- | --- | --- | --- | --- | --- |
|  | Estimate | P-value | 95% CI | Estimate | P-value | 95% CI |
| $\beta_0$ | 0.405 | <0.0001 | 0.346-0.463 | 0.405 | <.0001 | 0.346-0.463 |
| $\beta_1$ | 0.002 | 0.585 | -0.006-0.010 | 0.002 | 0.585 | -0.006-0.01 |
| $\beta_2$ | -0.087 | 0.010 | -0.148-(-0.025) | -0.087 | 0.010 | -0.148-(-0.025) |
| $\beta_3$ | 0.015 | 0.009 | 0.004- 0.025 | 0.015 | 0.009 | 0.004-0.025 |
| $\beta_1+\beta_3$ | 0.017 | <0.0001 | 0.012- 0.022 | 0.017 | <.0001 | 0.012-0.022 |
| **Male** |  |  |  |  |  |  |
| $\beta_0$ | 0.411 | <0.0001 | 0.345-0.477 | 0.043 | <.0001 | 0.036-0.050 |
| $\beta_1$ | 0.004 | 0.404 | -0.005-0.013 | 0.0004 | 0.317 | -0.0004-0.001 |
| $\beta_2$ | -0.10 | 0.011 | -0.173-(-0.028) | 0.0006 | 0.871 | -0.006-0.007 |
| $\beta_3$ | 0.015 | 0.019 | 0.003-0.027 | -0.0004 | 0.379 | -0.001-0.001 |
| $\beta_1+\beta_3$ | 0.019 | <0.0001 | 0.013-0.025 | 0.00001 | 0.948 | -0.0004-0.0004 |
| **Female** |  |  |  |  |  |  |
| $\beta_0$ | 0.39 | <0.0001 | 0.347-0.444 | 0.040 | <.0001 | 0.037-0.042 |
| $\beta_1$ | 0.0005 | 0.883 | -0.006-0.007 | 0.0002 | 0.314 | -0.0002-0.001 |
| $\beta_2$ | -0.07 | 0.012 | -0.121-(-0.019) | -0.001 | 0.658 | -0.007-0.004 |
| $\beta_3$ | 0.015 | 0.002 | 0.007-0.024 | 0.0002 | 0.455 | -0.0003-0.001 |
| $\beta_1+\beta_3$ | 0.016 | <0.0001 | 0.012-0.019 | 0.0004 | 0.083 | -0.0001-0.001 |
| **Age 18-64** |  |  |  |  |  |  |
| $\beta_0$ | 0.323 | <.0001 | 0.278-0.368 | 0.018 | <.0001 | 0.016-0.020 |
| $\beta_1$ | 0.006 | 0.0637 | -0.0001-0.012 | 0.0008 | <.0001 | 0.001-0.001 |
| $\beta_2$ | -0.049 | 0.0614 | -0.098-0.0003 | -0.001 | 0.511 | -0.005-0.002 |
| $\beta_3$ | 0.010 | 0.0235 | 0.002-0.018 | -0.0007 | 0.006 | -0.001-(-0.0002) |
| $\beta_1+\beta_3$ | 0.016 | <.0001 | 0.012-0.019 | 0.0002 | 0.209 | -0.0001-0.0004 |
| **Age => 65** |  |  |  |  |  |  |
| $\beta_0$ | 1.043 | <.0001 | 0.885-1.201 | 0.156 | <.0001 | 0.136-0.176 |
| $\beta_1$ | -0.008 | 0.456 | -0.029-0.013 | -0.001 | 0.400 | -0.004-0.002 |
| $\beta_2$ | -0.272 | 0.003 | -0.435-(-0.108) | -0.001 | 0.915 | -0.024-0.022 |
| $\beta_3$ | 0.046 | 0.003 | 0.019-0.074 | 0.002 | 0.212 | -0.001-0.005 |
| $\beta_1+\beta_3$ | 0.038 | <.0001 | 0.025-0.051 | 0.0009 | 0.243 | -0.001-0.002 |

$\beta_0$ : Constant value; $\beta_1$ : Pre- Trend slop; $\beta_2$ : Post- Level Change; $\beta_3$ : Post- Trend Change; $\beta_1+\beta_3$ : Post- Trend slop; CI: 95% Confidence interval; DPP4: Dipeptidyl Peptidase-4 Inhibitors

**Supplementary Table 3b:** Single ITS analysis (SITSA) for cumulative incidence rate of Metformin and Insulin before and after the pandemic in Sweden.

|  | **Metformin** |  |  | **Insulin** |  |  |
| --- | --- | --- | --- | --- | --- | --- |
|  | **Estimate** | **P-value** | **95% CI** | **Estimate** | **P-value** | **95% CI** |
| $\beta_0$ | 0.417 | <.0001 | 0.362-0.471 | 0.068 | <.0001 | 0.064-0.071 |
| $\beta_1$ | -0.004 | 0.268 | -0.011-0.003 | -0.0005 | 0.038 | -0.001-(-0.0001) |
| $\beta_2$ | -0.036 | 0.224 | -0.093-0.021 | 0.002 | 0.609 | -0.006-0.011 |
| $\beta_3$ | 0.007 | 0.083 | -0.0007-0.015 | 0.001 | 0.009 | 0.0003-0.007 |
| $\beta_1+\beta_3$ | 0.003 | 0.046 | 0.00006-0.006 | 0.0004 | 0.156 | -0.0002-0.001 |
| **Male** |  |  |  |  |  |  |
| $\beta_0$ | 0.454 | <.0001 | 0.392-0.515 | 0.074 | <.0001 | 0.066-0.082 |
| $\beta_1$ | -0.005 | 0.272 | -0.013-0.003 | -0.0009 | 0.117 | -0.002-0.0002 |
| $\beta_2$ | -0.039 | 0.256 | -0.104-0.027 | 0.004 | 0.576 | -0.010-0.018 |
| $\beta_3$ | 0.007 | 0.132 | -0.002-0.016 | 0.001 | 0.120 | -0.0002-0.002 |
| $\beta_1+\beta_3$ | 0.003 | 0.157 | -0.001-0.006 | 0.00006 | 0.884 | -0.001-0.001 |
| **Female** |  |  |  |  |  |  |
| $\beta_0$ | 0.385 | <.0001 | 0.339-0.431 | 0.065 | <.0001 | 0.063-0.068 |
| $\beta_1$ | -0.004 | 0.192 | -0.010-0.002 | -0.0004 | 0.017 | -0.0008-(-0.0001) |
| $\beta_2$ | -0.032 | 0.194 | -0.081-0.015 | 0.002 | 0.570 | -0.004-0.008 |
| $\beta_3$ | 0.008 | 0.021 | 0.002-0.015 | 0.001 | 0.001 | 0.0005-0.002 |
| $\beta_1+\beta_3$ | 0.004 | 0.002 | 0.001-0.007 | 0.0006 | 0.015 | 0.0001-0.001 |
| **Age 18-64** |  |  |  |  |  |  |
| $\beta_0$ | 0.372 | <.0001 | 0.329-0.415 | 0.061 | <.0001 | 0.057-0.065 |
| $\beta_1$ | -0.002 | 0.561 | -0.007-0.004 | -0.00003 | 0.890 | -0.001-0.0004 |
| $\beta_2$ | -0.012 | 0.625 | -0.059-0.035 | 0.002 | 0.711 | -0.007-0.011 |
| $\beta_3$ | 0.005 | 0.169 | -0.002-0.011 | 0.0008 | 0.070 | -0.00003-0.002 |
| $\beta_1+\beta_3$ | 0.003 | 0.032 | 0.0002-0.006 | 0.0008 | 0.039 | 0.00004-0.001 |
| **Age => 65** |  |  |  |  |  |  |
| $\beta_0$ | 0.991 | <.0001 | 0.852-1.131 | 0.138 | <.0001 | 0.123-0.153 |
| $\beta_1$ | -0.017 | 0.077 | -0.035-0.001 | -0.003 | 0.010 | -0.005-(-0.0008) |
| $\beta_2$ | -0.141 | 0.070 | -0.287-0.006 | 0.002 | 0.898 | -0.022-0.025 |
| $\beta_3$ | 0.025 | 0.020 | 0.005-0.045 | 0.003 | 0.003 | 0.001-0.006 |
| $\beta_1+\beta_3$ | 0.008 | 0.043 | 0.0002-0.016 | 0.0006 | 0.356 | -0.001-0.002 |

$\beta_0$ : Constant value; $\beta_1$ : Pre- Trend slop; $\beta_2$ : Post- Level Change; $\beta_3$ : Post- Trend Change; $\beta_1+\beta_3$ : Post- Trend slop; CI: 95% Confidence interval;

**Supplementary Table 3c:** Single ITS analysis (SITSA) for cumulative incidence rate of GLP1 agonist and SGLT2 inhibitors initiation before and after the pandemic in Sweden.

| <u>GLP1a</u> |  |  |  | <u>SGLT2i</u> |  |  |
| --- | --- | --- | --- | --- | --- | --- |
|  | <u>Estimate</u> | <u>P-value</u> | <u>95%CI</u> | <u>Estimate</u> | <u>P-value</u> | <u>95%CI</u> |
| $\beta_0$ | 0.029 | <.0001 | 0.024-0.033 | 0.020 | <.0001 | 0.016-0.024 |
| $\beta_1$ | 0.001 | 0.0001 | 0.0008-0.002 | 0.002 | <.0001 | 0.002-0.003 |
| $\beta_2$ | -0.008 | 0.014 | -0.015-(-0.002) | -0.033 | 0.003 | -0.054-(-0.013) |
| $\beta_3$ | 0.002 | <.0001 | 0.001-0.003 | 0.005 | 0.000 | 0.003-0.007 |
| $\beta_1+\beta_3$ | 0.003 | <.0001 | 0.003-0.004 | 0.007 | <.0001 | 0.009-0.009 |
| **<u>Male</u>** |  |  |  |  |  |  |
| $\beta_0$ | 0.024 | <.0001 | 0.018-0.031 | 0.016 | 0.0004 | 0.008-0.024 |
| $\beta_1$ | 0.001 | 0.002 | 0.0006-0.002 | 0.004 | <.0001 | 0.003-0.005 |
| $\beta_2$ | -0.010 | 0.023 | -0.018-(-0.002) | -0.051 | 0.002 | -0.079-(-0.022) |
| $\beta_3$ | 0.002 | 0.004 | 0.0007-0.003 | 0.006 | 0.003 | 0.002-0.009 |
| $\beta_1+\beta_3$ | 0.003 | <.0001 | 0.003-0.004 | 0.010 | <.0001 | 0.007-0.013 |
| **<u>Female</u>** |  |  |  |  |  |  |
| $\beta_0$ | 0.034 | <.0001 | 0.030-0.039 | 0.015 | <.0001 | 0.010-0.020 |
| $\beta_1$ | 0.001 | 0.001 | 0.0005-0.002 | 0.001 | 0.0002 | 0.0008-0.002 |
| $\beta_2$ | -0.006 | 0.073 | -0.013-0.0003 | -0.021 | 0.010 | -0.036-(-0.006) |
| $\beta_3$ | 0.003 | <.0001 | 0.002-0.004 | 0.003 | 0.005 | 0.001-0.005 |
| $\beta_1+\beta_3$ | 0.004 | <.0001 | 0.003-0.004 | 0.004 | <.0001 | 0.003-0.006 |
| **<u>Age 18-64</u>** |  |  |  |  |  |  |
| $\beta_0$ | 0.034 | <.0001 | 0.027-0.041 | 0.011 | <.0001 | 0.007-0.015 |
| $\beta_1$ | 0.001 | 0.005 | 0.0006-0.002 | 0.003 | <.0001 | 0.002-0.003 |
| $\beta_2$ | -0.005 | 0.271 | -0.014-0.004 | -0.015 | 0.004 | -0.024-(-0.005) |
| $\beta_3$ | 0.003 | <.0001 | 0.0019-0.005 | 0.001 | 0.072 | -0.00006-0.002 |
| $\beta_1+\beta_3$ | 0.005 | <.0001 | 0.004-0.006 | 0.004 | <.0001 | 0.003-0.005 |
| **<u>Age => 65</u>** |  |  |  |  |  |  |
| $\beta_0$ | 0.052 | <.0001 | 0.042-0.063 | 0.041 | <.0001 | 0.027-0.056 |
| $\beta_1$ | 0.0006 | 0.393 | -0.0008-0.002 | 0.005 | <.0001 | 0.003-0.007 |
| $\beta_2$ | -0.013 | 0.110 | -0.028-0.002 | -0.122 | 0.006 | -0.202-(-0.042) |
| $\beta_3$ | 0.002 | 0.006 | 0.0008-0.004 | 0.017 | 0.001 | 0.008-0.025 |
| $\beta_1+\beta_3$ | 0.003 | <.0001 | 0.002-0.004 | 0.022 | <.0001 | 0.014-0.030 |

$\beta_0$ : Constant value; $\beta_1$ : Pre- Trend slop; $\beta_2$ : Post- Level Change; $\beta_3$ : Post- Trend Change; $\beta_1+\beta_3$ : Post- Trend slop; SGLT2 inhibitor: Sodium-glucose co-transporter 2 inhibitors; CI: 95% Confidence interval; GLP1: glucagon-like peptide 1 agonists

## Notes

### Competing Interest Statement

The authors have declared no competing interest.

### Funding Statement

Yes

### Author Declarations

The research has ethics approval from the Swedish Ethical Review Authority (2020-01800 with subsequent amendments)

